# Fecal Short Chain Fatty Acid ratios are related to both Depressive and Gastrointestinal Symptoms in Young Adults

**DOI:** 10.1101/2020.04.24.20078352

**Authors:** Bettina Müller, Annica J. Rasmusson, David Just, Shishanthi Jayarathna, Ali Moazzami, Zorana Kurbalija Novicic, Janet L. Cunningham

**Author notes:** **Corresponding author:** Assoc. Prof. Janet Cunningham, Department of Neurosciences, Psychiatry, Uppsala University Hospital, SE-75185 Uppsala, Sweden, https://www.neuro.uu.se/research/research-groups/psychiatry/.

## Abstract

**Objective:** Short chain fatty acids (SCFAs) are produced by the gut microbiota and may reflect health. Gut symptoms are common in individuals with depressive disorders and recent data indicates relationships between gut microbiota and psychiatric health. We aimed to investigate potential associations between SCFAs and self-reported depressive and gut symptoms in young adults.

**Method:** Fecal samples from 164 individuals, of which 125 were patients with psychiatric disorders, were analyzed for the short chain fatty acids (SCFA) acetate, butyrate and propionate by nuclear magnetic resonance (NMR) spectroscopy. We then compared SCFA ratios to dimensional measures of self-reported depressive and gut symptoms.

**Results:** Depressive symptoms showed a positive association to acetate levels and negative associations to both butyrate and propionate levels in relation to total SCFA levels. Furthermore, symptoms of diarrhea and bloating showed positive associations to acetate and negative associations to propionate in relation to total SCFA levels. Cluster analysis revealed a heterogeneous pattern where shifts in SCFA ratios were seen for individuals with either elevated levels of depressive symptoms, elevated levels of gut symptoms or both.

**Conclusion:** Shifts in SCFAs may have relevance for both depressive symptoms and gut symptoms in young adults.

## Introduction

The World Health Organization (WHO) lifts Major Depressive Disorder (MDD) as a leading cause of disability in the world and it is currently associated with an annual suicide rate of approximately 800.000 individuals [1]. Furthermore, MDD and related disorders affects about 300 million people worldwide [2]. Yet, knowledge of the underlying pathophysiology is lacking, resulting in poor treatment and outcome for many patients [3].

It is clear that several neurotransmitter systems, including serotonergic, dopaminergic and glutamatergic systems may have altered function in MDD. Further, HPA (hypothalamic-pituitary-adrenal) axis abnormalities are described [4]. Previous studies have shown the potential influence of the gut microbiota and its effects on central regulatory systems that may influence body and brain function [5] including the HPA axis [6].

Short chain fatty acids (SCFA’s) are bacterial fermentation products that have the potential to influence the intestinal immune responses [7]. In the gut, the most abundant SCFAs are acetate, propionate and butyrate in a molar ratio of 3:1:1 accounting for >95% of the SCFA content [8]. Acetate, propionate and butyrate have multiple functions including energy supply, stimulation of epithelial cell proliferation, activation of G-protein coupled cell surface receptors as well as direct effects on substrate and energy metabolism in peripheral tissues such as the liver and skeletal muscles [8].

A new field of research indicates that bacterial products including SCFAs may also play a major role in stress-induced disorders including anxiety- and depressive-like behavior. A recent rodent study showed that SCFA supplementation could counter the effects of chronic psychosocial stress on anxiety- and depressive-like symptoms and behavior [9]. However, most of the studies investigating the role of SCFAs and their potential beneficial functions on brain and behavior are based on rodent models increasing the need of human studies on this topic. There are numerous reports linking irritable bowel syndrome (IBS) with depressive/anxiety disorders [10]. Since microbial dysbiosis has been implicated as causative to gut related symptoms in IBS, it is possible that shifts in SCFAs will be associated to both depressive symptoms and IBS.

The aim of our study was to test if the levels of acetate, propionate and butyrate are related to self-reported symptoms of depression and gastrointestinal symptoms in young adults with and without psychiatric disorders.

## Material and methods

### Ethical approval

The study was approved by the Regional Ethics Committee in Uppsala 2012/081 and 2013/219. All participants signed an informed consent.

### Participants

Participants in this study were patients recruited to participate in “Uppsala Psychiatric Patient Samples” (UPP), a project for collection of biological material from patients seeking psychiatric care at the Department of General Psychiatry at Uppsala University Hospital, Sweden. The patients in this study were recruited to UPP between September 2012 and February 2015. During this period, 388 patients were recruited to the biobank [11], and fecal samples from these 125 patients were available for SCFA analysis. Moreover, samples from 39 control individuals were available for SCFA analysis. The controls included staff members of the Uppsala University Hospital and students from Uppsala University. Assessment of psychiatric diagnoses was performed by trained personnel using the Structural Clinical Interview for DSM IV axis I disorders (SCID-I) or the Swedish version of the Mini-International Neuropsychiatric Interview (MINI).

### Depressive symptoms (MADRS-S)

The self-rating version of the Montgomery-Åsberg Depression Rating Scale – Self-Assessment (MADRS-S) was used for rating depressive symptoms [12-14]. The MADRS-S has been demonstrated to be a reliable patient-administered tool for depressive symptoms and consists of nine questions rated on a zero to six (0-6) Likert-scale, with a possible score range from zero to 54 [15].

### Gastro intestinal symptoms

We used the Swedish translation of the Gastrointestinal Symptom Rating Scale for IBS (GSRS-IBS) to evaluate gastro-intestinal symptoms. This is a validated self-assessment instrument for evaluating IBS symptoms, that is able to discriminate symptom severity and frequency [16]. The participants (both patients and controls) reported gastrointestinal (GI)-symptoms during the last week and scored the symptoms from one to seven (1-7) Likert-scale, with possible total scores ranging from 13 to 91 points. The questionnaire includes 13 questions that are grouped into symptom clusters; pain syndrome, bloating syndrome, constipation syndrome, diarrhea syndrome and satiety.

### Analysis of short chain fatty acids (SCFA)

SCFA has been analyzed using nuclear magnetic resonance (NMR) spectroscopy. Between 100-500 mg stool sample was diluted with 1.5 mL sodium phosphate buffer (0.4 M, pH 7.0), homogenized by vortex mixing and centrifuged at 6300 rpm at 4 °C for 15 min. From the supernatant, 1.5 mL of fecal water has been transferred to a fresh tube and subjected to centrifugation at 20 000 ×g at 4 °C for 15 min. This step was repeated once with 1 mL of supernatant recovered from the previous centrifugation. Finally, 525 µL of the supernatant was mixed with 45 µL D2O, and 30 µL internal standard. Each sample solution (560 µL) was transferred to a 5 mm NMR tube, and the 1H NMR spectra were acquired using a Bruker Advance III spectrometer operating at 600 MHz proton frequency and equipped with a cryogenically cooled probe and an auto sampler. Each spectrum was recorded (25 °C, 128 transients, acquisition time 1.8 s, relaxation delay 4 s) using a zgesgp pulse sequence (Bruker Biospin) using excitation sculpting with gradients for suppression of the water resonance. For each spectrum, 65 536 data points were collected over a spectral width of 17 942 Hz [18]. All NMR spectra were processed using Bruker TopSpin 4.1 software. The data were Fourier-transformed after multiplication by a line broadening of 0.3 Hz and referenced to internal standard peak TSP at 0.0 ppm. Baseline and phase were corrected manually. Each spectrum was integrated using Amix 3.7.3 (Bruker BioSpin GmbH, Rheinstetten, Germany) into 0.01-ppm integral regions between 0.41 and 10 ppm, in which areas between 4.52–5.00 ppm were excluded. Each integral region was referenced to the internal standard. The integral regions corresponding to short chain fatty acids were adjusted for mg of stool samples extracted and used for further statistical analysis. Few samples were analyzed with lesser sample amounts. Absolute numbers and calculation can be found in the supplementary file. Relative ratios of SCFAs were used for statistical analysis as these were less dependent on time or temperature differences during sample handling (preprint Cunningham, JL et al 2020) [17].

### Statistical analysis

The statistical correlation analyses were conducted with the Statistical Package for the Social Sciences (SPSS) Statistics Version 25. We calculated acetate, butyrate and propionate reference values as a ratio each to the total SCFAs (sum of acetate, butyrate and propionate) in the sample. Prior to all analyses, the variables were screened for distribution of normality with Shapiro-Wilk test of normality, *p*>0.05 and visually estimated for normal distribution (histogram, Q-Q-plot box-plot). We performed Mann-Whitney tests to investigate statistical difference between patient and control levels of acetate, butyrate and propionate, effect of gender and psychiatric medication. Bivariate analysis was performed to test if BMI and age were associated to SCFAs with Spearman’s rank correlation.

Correlation analysis (partial Spearman rank order correlations) were performed to analyze if acetate, butyrate and propionate ratios were related to depressive symptoms severity (MADRS-S total score) and self-reported gastrointestinal symptoms (GSRS-IBS). In all partial correlations analyses we controlled for body mass index (BMI) and use of psychiatric medication (yes/no). Individuals with incomplete or missing symptomatic data were also excluded from respective statistical analyses (Missing data for MADRS-S (n=2), GSRS-IBS (n=20), BMI n=5). The significance level was set to a *p*-value below 0.05 and since we considered our study as an extensive pilot study, with low statistical power, we did not perform any corrections for multiple testing. We did dimensional analyses and combined cases and controls, since controls were also displaying a range of both depressive and gut related symptoms.

Cluster analysis was performed using the statistical software R. A heat map was created to visualize the relationship between individual gut and depressive symptoms and the SCFA ratios (using the package *ggplots2*). For the cluster analysis, only 137 individuals had complete data from GSRS-IBS, MADRS-S and SCFA ratios which were respectively transformed to Z-scores. To visualize the distribution of data, high levels or scores are coded orange while low levels or scores are coded in purple. The BMI on the left side of the heat map is colored blue, where low BMI is colored light blue. To illustrate patients and controls, on the right-side values that represent patients are in black and controls are marked grey. Furthermore, to visualize the clusters better, we colored the columns showing MADRS-S items in pink, SCFA ratios in green and the GSRS-IBS items in turquoise.

## Results

The cohort’s characteristics are described in Table 1. In the bivariate analyses, acetate ratios were negatively associated to BMI (-0.156, p=0.049), but no associations were found to butyrate ratios (0.103, p=0.196) nor propionate ratios (0.132, p=0.097). No relationship between age or gender to SCFA ratios were found. Patients did not differ as a group from controls regarding acetate, butyrate and propionate ratios (Supplementary Table 1). However, we could see a tendency towards an association between acetate and butyrate ratios and psychiatric medication and use of anti-depressants (Supplementary Table 1). Based on these findings, the partial correlations below were adjusted for BMI and psychiatric medication (Y/N).

**Table 1:** Relevant characteristics of the cohort

|  |  | Patients (n=125) | Controls (n=39) |
| --- | --- | --- | --- |
| Age (mean ± SD), years | | 21.9 (2.6) | 28.5 (9.5) |
| Gender (Men/Women) % Men |  | 17/108 (14%) | 15/24 (38%) |
| BMI (median, range) |  | 22.4 (15.1-46.6) | 23.0 (18.1-37.8) |
| MADRS-S total score (median, range) |  | 22 (0-49) | 5 (0-20) |
| GSRS-IBS total score (median, range) |  | 31.5 (13-78) | 23.0 (13-47) |
| Medication N (%) | Any medication N (%) | 106 (84.8%) | 20 (51.2) |
|  | Any psychiatric medication N (%) | 82 (66%) | 3 (8%) |
|  | Other anxiolytic medication N (%) <sup>a</sup> | 29 (23%) | 0 (0%) |
|  | Anti-depressive treatment N (%) <sup>b</sup> | 58 (46%) | 3 (7.7%) |
|  | Antipsychotics N (%) | 7 (5.6%) | 0 (0%) |
|  | Anti-epileptics N (%) | 11 (8.8%) | 0 (0%) |
|  | Benzodiazepines N (%) | 4 (3.2%) | 0 (0%) |
|  | Z-analogues N (%) | 7 (5.6%) | 0 (0%) |
|  | Immune modulating N (%) | 8 (6.4%) | 0 (0%) |
| Diagnosis N (%) | Current depressive episode(UP/BP) | 68 (54.4%) | 0 (0%) |
|  | Any bipolar disorder <sup>c</sup> | 23 (18.4%) | 0 (0%) |
|  | Celiac disease | 1 (0.8%) | 0 (0%) |
|  | Systemic inflammatory disease <sup>d</sup> | 4 (3.2%) | 0 (0%) |
|  | Lifetime depressive episode (UP) | 73 (58.4%) | 5 (13.8%) |
|  | Any anxiety disorder DSM-IV | 85 (68%) | 1 (2.5%) |
|  | Anorexia | 0 (0%) | 0 (0%) |
|  | Bulimia | 10 (8%) | 0 (0%) |
| SCFAs | Acetate ratio, median (range) | 0.77 (0.45-0.98) | 0.76(0.44-0.87) |
|  | Butyrate ratio, median (range) | 0.05 (0.01-0.12) | 0.05 (0.03-0.09) |
|  | Propionate ratio, median (range) | 0.18 (0.01-0.43) | 0.18 (0.08-0.51) |

*Abbreviations:* BMI = body mass index; missing data for 4 patients, MADRS-S = Montgomery-Åsberg Depression Rating Scale-Self-Assessment; missing data for 1 patient, GSRS-IBS = Gastrointestinal Symptom Rating Scale; missing data for 14 patients and 4 controls, UP/BP = Unipolar/Bipolar disorder, SSRI = Selective serotonin reuptake inhibitors, SNRI = serotonin-norepinephrine reuptake Inhibitor. a - Sedating antihistamines, phenothiazines. b - SSRI, SNRI, mood stabilizers and atypical antidepressants. c - Bipolar type I and II and non-specified - Systemic inflammatory disease *i.e.* four cases of inflammatory bowel disorder (IBD). One case of Asperger's and dyslexia, one case of Attention deficit hyperactivity disorder (ADHD), one case presented psychotic symptoms, and one patient has since this study started committed suicide. SCFAs = short chain fatty acids.

Depressive symptoms severity (total MADRS-S score) was correlated with acetate ratio (r=0.235, p=0.003), butyrate ratio (r= -0.195, p=0.014) and propionate ratio (r= -0.207, p=0.009) when adjusting for BMI and psychiatric medication.

Exploratory analyses to investigate the associations to the separate MADRS-S items (9 questions) and the three SCFA ratios were performed in a post-hoc analysis (shown in Table 2). Positive associations were found between acetate ratio and five of the MADRS-S items (mood, feelings of unease, initiative, emotional involvement, and pessimism), but no significant associations were found for sleep, ability to concentrate, appetite and zest for life. Butyrate and propionate showed an inverse pattern of associations to the same items as acetate (Table 2).

**Table 2.** Relation of depressive symptoms and acetate, butyrate and propionate ratios, shown by correlation coefficients and 2-tailed *p*-values for partial bivariate associations between the SCFAs ratios and the Montgomery-Åsberg Depression Rating Scale – Self-Assessment (MADRS-S) and the separate items. Adjusted for BMI, age and use of psychiatric medications (Y/N). Bold values indicate an association with a *p*-value of less than 0.05. The analysis between SCFAs and MADRS-S total score is the main hypothesis of our study, and the individual items are exploratory and thus not corrected for multiple testing.

| *Main hypothesis (n=155)* | Acetate | Butyrate | Propionate |
| --- | --- | --- | --- |
| **MADRS-S Total score** | **0.235 (0.003)** | **-0.195 (0.014)** | **-0.2017 (0.009)** |
| *Post Hoc* |  |  |  |
| **MADRS-S individual items** |  |  |  |
| Mood | **0.227 (0.006)** | **-0.198 (0.018)** | **-0.199 (0.017)** |
| Feelings of unease | **0.184 (0.028)** | -0.074 (n.s.) | **-0.185 (0.026)** |
| Sleep | 0.054 (n.s.) | 0.020 (n.s.) | -0.065 (n.s.) |
| Appetite | 0.114 (n.s.) | -0.088 (n.s.) | -0.102 (n.s.) |
| Ability to concentrate | 0.152 (n.s.) | -0.139 (n.s.) | -0.131 (n.s.) |
| Initiative | **0.227 (0.006)** | **-0.203 (0.002)** | **-0.198 (0.017)** |
| Emotional involvement | **0.202 (0.015)** | **-0.174 (0.037)** | **-0.177 (0.034)** |
| Pessimism | **0.208 (0.012)** | **-0.163 (0.050)** | **-0.187 (0.024)** |
| Zest for life | 0.159 (n.s.) | -0.096 (n.s.) | -0.151 (n.s.) |

The subsequent exploratory analysis of correlations between SCFA ratios and diverse gastrointestinal symptom items (GSRS-IBS) are shown in Table 3. A positive association was found between acetate ratios and the GSRS-IBS items diarrhea and bloating, while negative associations were found between constipation and butyrate ratios and diarrhea and propionate ratios (Table 3.) These findings were not affected by adjustment factors of BMI and psychiatric medications (data not shown).

**Table 3.** Post Hoc analysis of acetate, butyrate and propionate ratios in relation to gastrointestinal symptoms measured using the Gastrointestinal Symptom Rating Scale for IBS (GSRS-IBS). Correlation coefficients and 2-tailed *p*-values in brackets for associations between SCFA ratios and the separate GSRS-IBS-items. N=152. Adjusted for BMI, age and use of psychiatric medications (Y/N). Bold values indicate an association with a *p*-value of less than 0.05.

| **GSRS-IBS Symptoms** | **Acetate** | **Butyrate** | **Propionate** |
| --- | --- | --- | --- |
| Satiety | 0.076 (n.s.) | -0.113 (n.s.) | -0.053 (n.s.) |
| Diarrhea | **0.217 (0.010)** | -0.048 (n.s.) | **-0.229 (0.007)** |
| Constipation | -0.031 (n.s.) | **-0.169 (0.046)** | 0.082 (n.s.) |
| Bloating | **0.172 (0.043)** | -0.156 (n.s.) | -0.148 (n.s.) |
| Pain | 0.117 (n.s.) | -0.120 (n.s.) | -0.098 (n.s.) |

To visualize the relationships between the symptoms and SCFA ratios, we performed cluster analyses. As shown in Figure 1, a heat-map illustrates the clusters of the separate items of MADRS-S and GSRS-IBS, as well as the acetate, butyrate and propionate ratios. (Figure 1). Six clusters are highlighted in the heat map, illustrated as black rectangles around the respective grouping. Cluster I contains patients with mixed depressive symptoms and gastrointestinal symptoms with predominance of diarrhea and lower propionate ratios. Cluster II includes patients with high depressive scores and lower scores on other gut symptoms and shows lower butyrate and propionate ratios. Group 3 (III) describes a small group of individuals with both low gut and depressive symptoms. Interestingly, this small group has very low acetate ratios and higher butyrate and propionate ratios. Cluster IV includes patients with both depressive symptoms and gut related issues with predominance of pain and obstipation and lower butyrate ratios. Cluster V includes controls and patients with some gastrointestinal symptoms and elevated acetate ratios. Cluster VI, in the lowest rectangle, controls with low depressive scores but with some gastrointestinal symptoms also appear to have lower acetate and propionate ratios and somewhat higher butyrate ratios.

**Figure 1.**
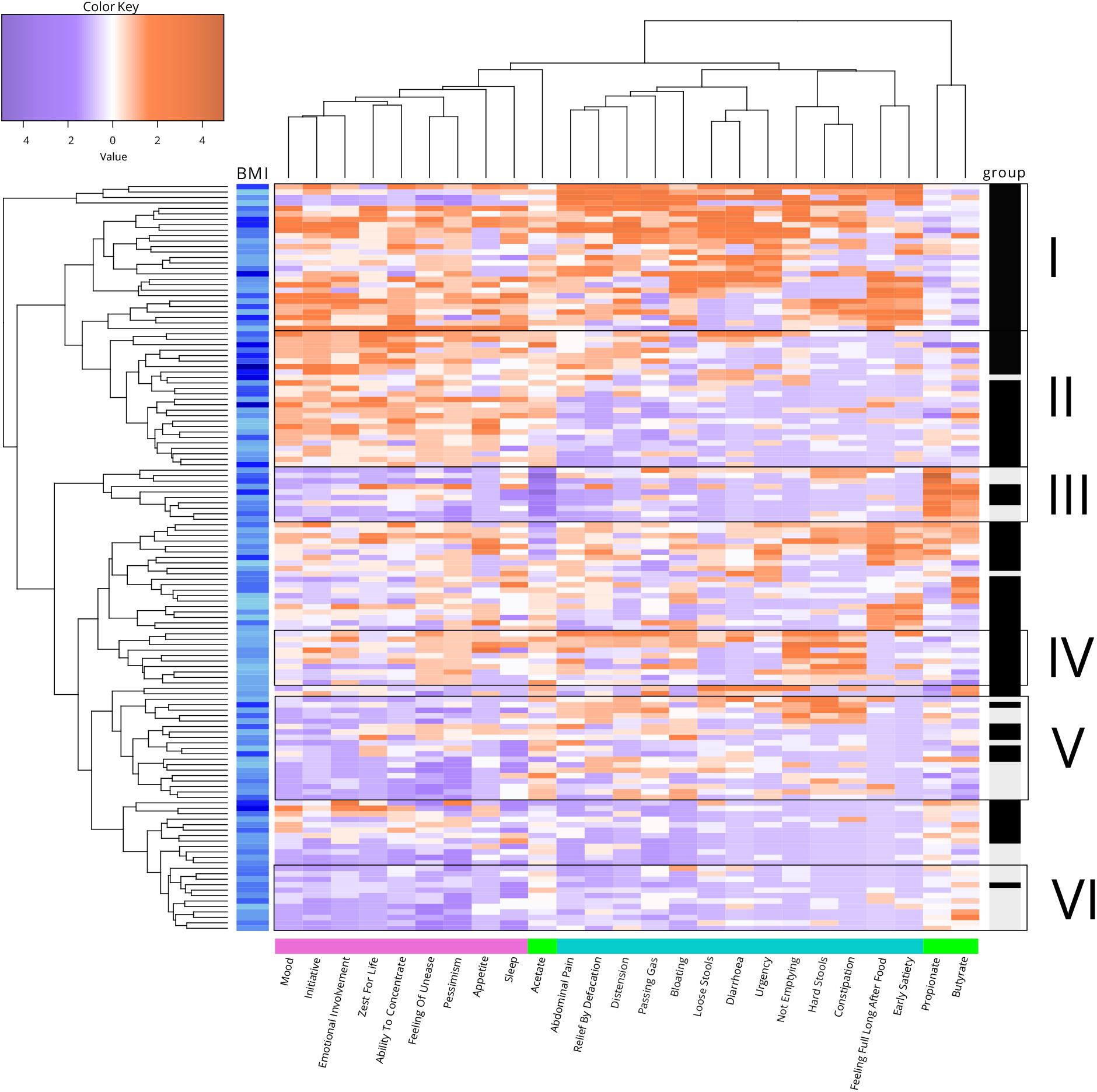
Heat-map of the separate MADRS-S and GSRS-IBS-items and acetate, butyrate and propionate ratios. A heat map visualizing the relationship between individual gut and depressive symptoms and SCFA ratios. For the cluster analysis, only 137 individuals had complete data from GSRS-IBS, MADRS-S and SCFA analysis. Orange boxes represent high levels/scores of the respective items and a purple represent low levels/scores of these items. BMI levels are indicated in blue, where light blue indicates low BMI (left panel). To illustrate individual groups, patients are colored with black boxes and controls in grey boxes (right panel). The MADRS-S items are in pink, SCFAs in green and the GSRS-IBS items in turquoise (lower panel).

## Discussion

This pilot study found relationships between depressive and gastrointestinal symptoms and the ratios of three major SCFAs measured in fecal material from a mixed cohort of young adults with and without psychiatric diagnoses. These associations were independent from BMI and psychiatric medication. Further, exploration of the data revealed groups of individuals with either depressive symptoms, gastrointestinal symptoms alone - or in combination - may show shifts in SCFA ratios with relevance for both patients and controls.

Our results indicate that shifts in SCFA ratios may contribute to both IBS and depression. A bidirectional interaction between gut symptoms and the brain mediated by the gut microbiota and their respective metabolites has been proposed [19-21]. SCFAs in addition to local influences on epithelial cells [22] activate G protein-coupled receptors [23] and are involved in neurotransmitter production [24]. Moreover, SCFAs can penetrate the blood-brain barrier [25]. Differences in the SCFA composition, because of changes in microbiota, correlates with neural activity and brain structure in humans, as assessed by functional and structural MRI [26].

While studies examining SCFAs differ in their design by measuring different populations, feces or blood and calculating absolute or relative levels which makes comparison between studies more difficult, some findings are consistent. Our study showed that acetate and butyrate are associated with depressive and gastrointestinal symptoms, but the relationships appears to go in opposite directions. In agreement with earlier work, acetate was positively correlated to BMI in our material. Elevated levels of acetate in feces has been shown to be associated with obesity in several studies and was recently summarized in a meta-analysis but does not reach significance after corrections for cofactors in other studies on older populations [27]. The relationship to depressive and gastrointestinal symptoms were independent from BMI in this young adult population raising an interesting hypothesis that these symptoms in combination with SCFA disruption may predispose for future metabolic syndrome and obesity in later life. The negative association between butyrate ratio and depressive gastrointestinal symptoms in this study was weak, and is in line with earlier findings and proposed positive impact of butyrate on health [28]. These findings raise questions as to the potential future therapies. Oral administration of butyrate has been suggested to be beneficial for a wide spectrum of diseases, ranging from metabolic diseases to colorectal cancers [29]. Fiber rich diets such as the “Mediterranean style” diet are also reported to increases butyrate and, in theory, may reduce the risk for several diseases linked to aging and the immune system [30]. As we can conclude from the clustering data, several groups of individuals with different combinations of SCFA distribution, depressive and or gastrointestinal symptoms exist. Whether symptoms in these groups respond to dietary or supplement-based interventions in a uniform or differential manner remains to be seen. Some indications of highly individual reactions to dietary interventions are emerging [31].

The present study has some limitations. The population is small and individuals with psychiatric morbidity are overrepresented. The management of samples during collection may have varied between individuals, however our previous data show that the use of SCFA ratios instead of absolute values reduced the influence of time and temperature on the results [17] (preprint Cunningham, JL et al 2020). Furthermore, we do not have information of dietary pattern or access to repeated sampling. A strength of the study is the use of dimensional measures of depressive and gastrointestinal symptoms and we have a full range of symptoms represented in the cohort.

This study demonstrates clear interactions between psychiatric and gastrointestinal symptoms with SCFA composition. Patients with psychiatric disease are a heterogeneous group; this study provides insight about potential subgroups with high acetate and low butyrate/propionate ratios that should be further explored regarding the potential beneficial effects after dietary inventions.

## Data Availability

All data are available in the supplementary file.

## Acknowledgements

The authors thank Ulla Nordén for her excellent research assistance, Hans Arinell for excellent statistical advice and Uppsala Biobank for collaboration in sample management.

## List of acronyms

SCFAs: Short chain fatty acids
WHO: World Health Organization
MDD: Major Depressive Disorder
HPA: hypothalamic-pituitary-adrenal
IBS: irritable bowel syndrome
UPP: Uppsala Psychiatric Patient Samples
SCID-I: Structural Clinical Interview for DSM IV axis I disorders
MINI: Mini-International Neuropsychiatric Interview
MADRS-S: Montgomery-Åsberg Depression Rating Scale – Self-Assessment
GSRS-IBS: Gastrointestinal Symptom Rating Scale for IBS
NMR: nuclear magnetic resonance
SPSS: Statistical Package for the Social Sciences
BMI: body mass index
MRI: Structural magnetic resonance imaging
UP/BP: Unipolar/Bipolar disorder
SSRI: Selective serotonin reuptake inhibitors
SNRI: serotonin-norepinephrine reuptake Inhibitor
ADHD: Attention deficit hyperactivity disorder
SD: standard deviation

---

**Supplementary Table 1.** Mann-Whitney independent samples test, *p*-values. *Abbreviations:* SSRI = Selective serotonin reuptake inhibitors, SNRI = serotonin-norepinephrine reuptake inhibitor

|  | Acetate | Butyrate | Propionate |
| --- | --- | --- | --- |
| Cases vs. controls | 0.333 | 0.776 | 0.211 |
| Gender | 0.340 | 0.654 | 0.229 |
| Psychiatric medication | 0.429 | 0.076 | 0.707 |
| Anti-depressive | 0.080 | 0.107 | 0.185 |
| SSRI | 0.067 | 0.113 | 0.206 |
| SNRI | 0.814 | 0.836 | 0.636 |
| Anxiolytics | 0.467 | 0.980 | 0.310 |

